# Seasonal variations and time trends of deaths from COVID-19 in Italy, September 2021 - September 2024: a segmented linear regression study

**DOI:** 10.1101/2025.01.25.25321115

**Authors:** Marco Roccetti, Eugenio Maria De Rosa

## Abstract

Using a segmented linear regression model, we examined the seasonal profiles of weekly COVID-19 deaths data, in Italy, over a three-year long period during which the SARS-CoV-2 Omicron and post-Omicron variants were predominant (September 2021 - September 2024). Comparing the slopes of the regression segments, we were able to discuss the variation in steepness of the Italian COVID-19 mortality trend, identifying the corresponding growth/decline profile for each considered season. Our findings show that, although the COVID-19 weekly death mortality has been in a declining trend in Italy since the end of 2021 until the end of 2024, there have been increasing alterations of the COVID-19 deaths for all winters and summers of that period. These increasing mortality variations were more pronounced in winters than in summers, with an average progressive increase of the number of COVID-19 deaths, with each new week, of 55.75 and 22.90, in winters and in summers, respectively. We found that COVID-19 deaths were, instead, less frequent in the intermediate periods between winters and summers, with an average decrease of -38.01 COVID-19 deaths for each new week. Our segmented regression model has fit well the observed COVID-19 deaths, as confirmed by the average value of the determination coefficients: 0.74, 0.63 and 0.70, respectively for winters, summers and intermediate periods, in a scale from 0 to 1. In conclusion, favored by a general declining COVID-19 mortality trend in Italy in the period of interest, transient rises of the mortality have occurred both in winters and in summers, but received little attention. The reason why these increasing alterations of the COVID-19 seasonal deaths have been considered cause for concern only occasionally is that they have been always compensated by consistent downward drifts occurring during the intermediate periods between winters and summers.

## 1. Introduction

The COVID-19 pandemic has become a subject of increasing and widespread concern for Italy, particularly with the Omicron variant and its swiftly mutating sub-lineages which stabilized as the predominant cause of infection for almost three years, corresponding to a period which extended from September 2021 to September 2024 [1]. Fortunately, a broad spectrum of timely response strategies (including vaccination) has resulted in a progressively reduced COVID-19 mortality [2], which has followed a declining trend, from an average number of weekly deaths of almost 1000 in the period October 2021 - September 2022 to around 100 in the period October 2023 - September 2024 [3, 4].

Despite the decision with which the World Health Organization (WHO), on May 5, 2023, announced the end of the emergency phase and the beginning of the COVID-19 post-pandemic era [5], the virus has not been eradicated, and with each new winter season the question remains if, like many other respiratory virus illnesses, COVID-19 could peak during the winter, favored by: new variants, decreasing immunity from previous infections and vaccinations, environmental conditions, human behaviors like dense people gathering, relaxation of public health measures for prevention and control [6, 7, 8]. The question above, about the possibility of COVID-19 following a one-year seasonal pattern similar to many other viral infections, has been intensively discussed by the scientific community. Two opposite sides have appeared quite clearly: on one side, those convinced of the existence of a seasonal pattern that repeats over a fixed one-year period (at least, in all the Western countries) [9]; on the other, those bringing to the table evidences of several repeating outbreaks, not necessarily occurring on a yearly basis [10]. We have had this discussion repeated many times, over these years, in Italy, with the two sides offering mostly the same arguments, with occasional country-specific remarks.

In this complex context, despite there is accumulating evidence against the hypothesis of a sinusoidal seasonality of the COVID-19 illness assumed to recur over a one-year period solely, an analysis of the time series of the COVID-19 deaths in Italy, over the period end of 2021 - end of 2024, has shown the presence of recurrent alterations in this mortality trend that are not simply attributable to occasional upward/downward drifts of the data time series [11]. This was the motivation for better investigating the occurrence of seasonal variations of COVID-19 mortality in Italy, over the period during which the initial Omicron variant diversified into multiple sub-variants that gained an extremely increased survival fitness, leading to pandemic waves recurring in different seasons of the same year.

To identify the seasonal profiles of deaths from COVID-19 in the period of interest, we adopted a mathematical approach resulting in a segmented linear regression model of the COVID-19 deaths data, where each increasing/decreasing seasonal death trend variation corresponds to a regression segment with a given steepness. Comparing the slopes of these regression segments, we have been able to discuss the alterations of the mortality trend, identifying the corresponding growth/decline profiles for each considered season. The utilized time series of COVID-19 deaths data is publicly available and it is provided by both the Italian Civil Protection Department and the Italian Ministry of Health, on a weekly basis.

This allowed us to identify non-occasional variations of the number of deaths from COVID-19, over the three-year long period of interest, with increasing alterations of the mortality trend present both in winters and in summers, but more pronounced in winters. In particular, the average progressive increase of the number of COVID-19 deaths, for each new week, was of 55.75 and 22.90, in winters and summers, respectively. COVID-19 deaths were, instead, less frequent in the intermediate periods between winters and summers, with an average decrease of -38.01 COVID-19 deaths, for each new week. The measure of how well our linear regression model has fitted the observed COVID19 deaths data is confirmed by the average values of the determination coefficients, returned by the model in the neighborhood of 70%. In addition, it should be also considered that all the *p-values* computed during the use of our model were below the significative value of *α* = 0.05, thus providing a further confirmation of the statistical validity of this analysis.

Before concluding this Section, it is also worth mentioning the following facts. First, to the best of our knowledge, this paper is the first to analyze a very specific epidemiological landscape, in terms of the COVID-19 mortality, characterized by a given geography (Italy) and a very long time period of observation where only the SARS-CoV-2 Omicron and post-Omicron sub-lineages were predominant (September 2021 - September 2024). Second, this is an observational study which has deliberately avoided the problem to quantify the role attributable to the various factors that can have had an influence on COVID-19 deaths, including prevention/control measures and vaccinations. Third, we recognise that the method we have proposed (segmented linear regression) is not the one with which to count COVID-19 deaths. Methods based on Poisson-like distributions would be more appropriate in that case. In fact, the target of this study was not trying to count the deaths from COVID-19 precisely per each single week of observation, but to look at how quickly they grew or decline, comparing the slopes of the regression segments. Finally, as to the health policy recommendations resulting from our research, we are confident we have provided an important contribution for the benefit of that subset of at risk population who are seasonally vulnerable, having clearly identified the seasons to consider with more attention.

The remainder of this paper is the following. In the Materials and Methods Section, we first describe where our data come from and how they have been temporally organized for our study, then we explain the methodology we have used to analyze them. In the Results Section, we describe the results we have obtained, and in the Discussion Section, we describe both the advantages and the limitations of our approach. Finally, the Conclusions Section terminates our paper.

## 2. Materials and methods

### 2.1. Sources of data and linear regression segments

In this section, we provide sufficient details to allow readers to replicate our results. We decided to work with the time series of the Italian COVID-19 deaths data, to which some simple transformations were applied as described in the following. We anticipate here that all the initial COVID-19 deaths data are downloadable from two public, open access repositories, specifically: i) the repository maintained by the Italian Civil Protection Department, under the Italian Presidency of the Council of Ministers (https://github.com/pcm-dpc/COVID-19/blob/master/dati-andamento-nazionale), and ii) the repository maintained by the Italian Ministry of Health, (https://www.salute.gov.it/new/it/tema/covid-19/report-settimanali-covid-19/). With this data, we fitted a segmented linear regression model [12], where the dependent variable was the number of weekly confirmed COVID-19 deaths, and the independent variable was the number of weeks since 23 September 2021 until 19 September 2024, totaling 157 weeks. The result has been a model comprised of a series of segments, each connecting two points, beginning at one and ending at the other. Unlike a continuous line, a regression segment is defined by these two points. Those couples of points were chosen based on two different criteria to discern an increasing variation of the deaths data time series from a decreasing one. For an increasing variation, the starting point was chosen in correspondence with the beginning of a given pandemic wave, while the corresponding end point was the point in time when that wave peaked. As to a decreasing variation, the starting point was chosen in correspondence with the weeks immediately subsequent to a peak, while the end point corresponded to the time when that wave returned to baseline values. Since micro oscillations of the number of weekly deaths (up to +/- 15%) are possible along a path of consecutive points of a deaths time series (both during an ascending and a descending phase), the rule above was not implemented strictly, sometime allowing oscillating, but near, points not to be interpreted as a definitive change of direction of the deaths trend, from increasing to decreasing, or vice versa. This occurs quite typically at the beginning of a wave or during the weeks after it has peaked.

### 2.2. COVID-19 deaths data time series

Given the existing literature that hypothesizes that winters and summers are the seasons when COVID-19 waves often occur [13, 14], we focused our attention on those two seasons. Taking into account the specificity of the Italian climate, we considered an extended definition of the winter season which also included high fall, with a corresponding timeframe extending from beginning of September to end of January.

As to summer, we considered a timeframe starting from end of May/beginning of June to end of August. Within the extension of these two timeframes, we looked for increasing trends of COVID-19 deaths data, each with a duration of at least six weeks (one month and a half). Choosing six weeks comes from the working definition of COVID-19 waves as provided in [15], where the three quarters of the upward trends of many studied COVID-19 waves lasted something more than a month. Similarly, for the downward trends.

With this initial analysis we identified three fall-winter periods (from now on, only wither for short) and three summer periods, where noticeable increasing trends of the COVID-19 mortality were observed. What remains of the COVID-19 deaths data time series, after that the six periods with ascending trends are removed, corresponds exactly to three periods where, not surprisingly, noticeable descending trends of the COVID-19 mortality time series can be identified. It is interesting to point out that all these three periods, with declining COVID-19 mortality profiles, are set in between the end of winters and before the beginning of the subsequent summers: a kind of extended spring periods. For that reason, we have indicated those periods with the term intermediate.

As a result of the preliminary procedures described above, the initial COVID-19 deaths data time series was divided over nine different periods, six of which with increasing mortality trends and three with decreasing trends. The first and the last week of each identified period represent, respectively, the starting and the end points of the regression segments we will try to fit with our regression model.

A graphical summary of the entire COVID-19 deaths data time series is plotted in Figure 1, where all the nine periods are represented with colored sectors to differentiate winters, summers and intermediate seasons. In particular, blue nuances are for winters, red nuances for summers and grey ones for intermediates.

**Figure 1.**
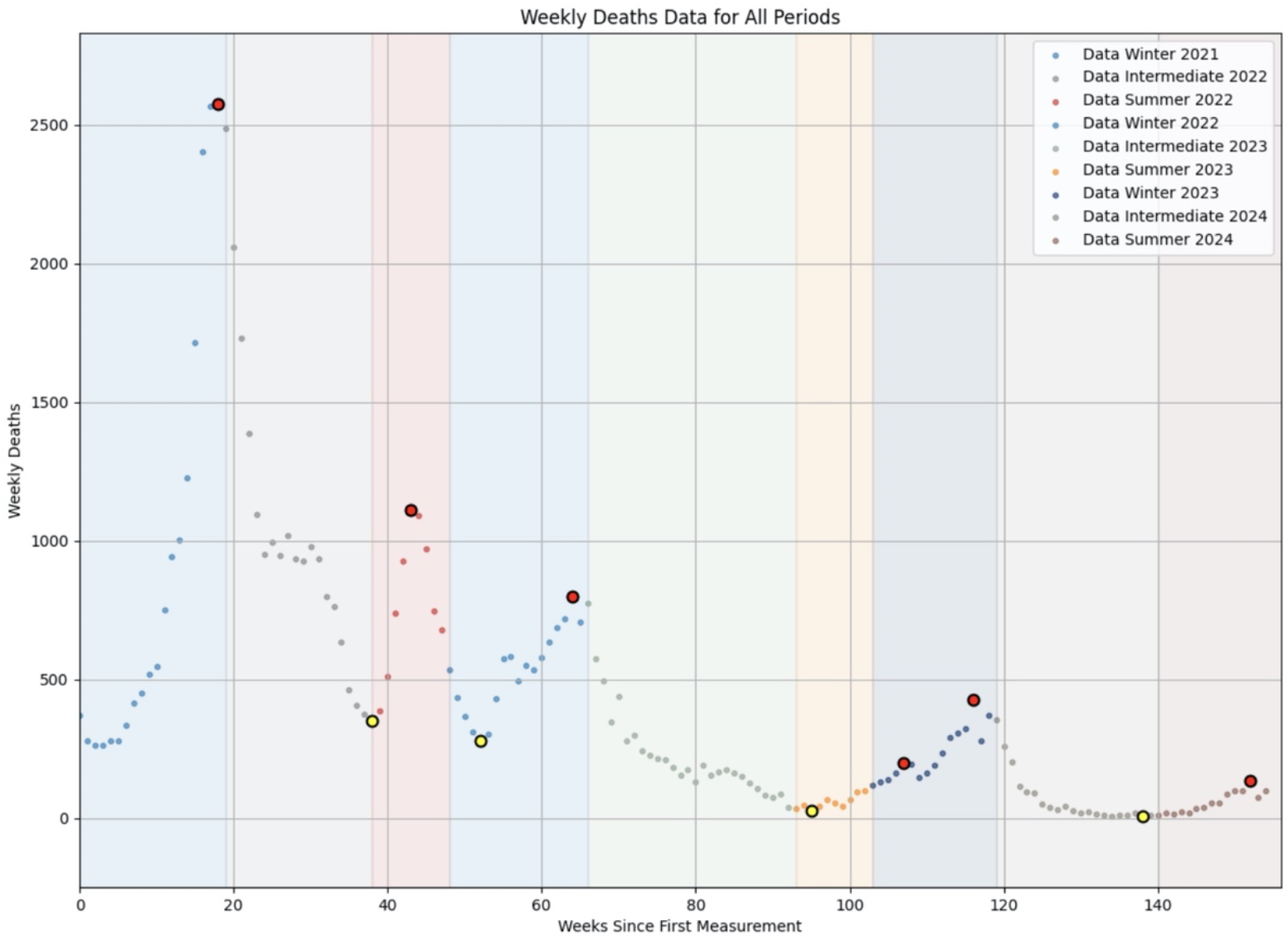
COVID-19 deaths data time series: 23 September 2021 - 19 September 19 2024. y axis: number of weekly deaths; x axis: weeks from 1 to 157. Colored sectors for nine different seasons with corresponding deaths. Blue nuances, fall-winter; red nuances, summer; grey nuances, intermediate periods. Big red dots for the six peaks of six ascending trends. Yellow dots for the main local minima.

The six peaks coming at the culmination of the six ascending trends mentioned above are put in evidence, in the Figure, with big red dots. They occurred in correspondence of the weeks ending with the following dates: January 28, 2022 (2,575 weekly deaths); July 7, 2022 (1,111 weekly deaths); December 16, 2022 (798 weekly deaths); October 12, 2022 (197 weekly deaths); December 14, 2023 (425 weekly deaths) and August 22, 2024 (135 weekly deaths). With big yellow dots, we have also marked the principal local minima of this death data time series.

In addition, Table 1 reports the main characteristics of the COVID-19 deaths data time series depicted in Figure 1. Among these, the dates are reported corresponding to the starting and the end points of each period of Figure 1, which will be be used to compute the segments of our regression model. To be noticed are also the mean durations in weeks of winter, summer and intermediate periods of Figure 1 which are, respectively equal to: 17.66 (Winter 2021, Winter 2022, Winter 2023), 12.33 (Summer 2022, Summer 2023, Summer 2024) and 22.33 (Intermediate 2022, Intermediate 2023, Intermediate 2024), with standard deviation (SD) values, respectively, of: 1.25, 3.30, 3.40 weeks. The number of deaths, divided over the three years of interest, is reported in the rightmost column of the Table (both cumulative and averaged per week).

**Table 1.** Characteristics of the COVID-19 deaths data time series: period, season type, increasing/decreasing mortality trend, number of weeks per period, dates of beginning/end points, number of deaths per year (cumulative and weekly average).

| Period | Season type | Deaths trend | Number of weeks | Beginning point of regression segment | End point of regression segment | Number of COVID-19 Deaths per year |
| --- | --- | --- | --- | --- | --- | --- |
| Winter 2021 | Fall-Winter | Increasing | 19 | Sep 23, 2021 | Jan 28, 2022 | 44,576 total, |
| Inter. 2022 | Extended Spring | Decreasing | 19 | Feb 4, 2022 | Jun 10, 2022 | 928,67 |
| Summer 2022 | Summer | Increasing | 10 | Jun 17, 2022 | Aug 19, 2022 | weekly avg./ |
| Winter 2022 | Fall-Winter | Increasing | 18 | Aug 26, 2022 | Dec 23, 2022 | 16,349 total, |
| Inter. 2023 | Extended Spring | Decreasing | 27 | Dec 30, 2022 | Jun 30, 2023 | 297,25 |
| Summer 2023 | Summer | Increasing | 10 | Jul 7, 2023 | Sep 7, 2023 | weekly avg./ |
| Winter 2023 | Fall-Winter | Increasing | 16 | Sep 14, 2023 | Dec 28, 2023 | 6,160 total |
| Inter. 2024 | Extended Spring | Decreasing | 21 | Jan 4, 2024 | May 23, 2024 | 114,07 |
| Summer 2024 | Summer | Increasing | 17 | May 30, 2024 | Sep 19, 2024 | weekly avg./ |

It is useful to remind, in the end of this discussion, that we are treating the concept of season as super-long (or even slightly reduced) periods of cold and warm, plus other characterizing climatic factors; not seasons in an astronomical sense with a strictly respect for when equinoxes and solstices occur.

### 2.3 Method of analysis

The segmented (or piecewise) regression model we used to fit the COVID-19 deaths data of Figure 1 follows the formula:

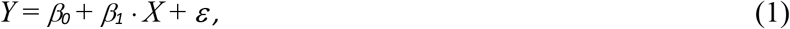

where *Y* corresponds the number of weekly COVID-19 deaths and *X* represents the passage of time measured in weeks. *β*_*0*_ is the intercept, that is the value of *Y* when *X* is equal to 0; *β*_*1*_ is the slope (or slope coefficient) of a regression segment and indicates the steepness of that segment. Finally, *ε* represent the error term [16].

In the specific case of our model, *β*_*1*_ indicates the rate, or the velocity, with which a segment reflects an increasing/decreasing mortality trend, while *β*_*0*_ registers the portion of *Y* (number of deaths) not influenced by *X* (passage of time), in some sense it shows how well the linear model approximates the general mortality situation prior to the beginning of a given increasing/decreasing mortality trend.

In our linear model *β*_*1*_ plays a major role as it informs about the change of the dependent variable *Y* (number of deaths) for a one week increase (*X*). In simple words, *β*_*1*_ represents how much the number of COVID-19 deaths has increased, with a one week increase in the passage of time. The larger *β*_*1*_, the steeper the slope of the segment, and correspondingly the speedier the increase of the number of deaths.

In essence, with our analysis we are trying to find estimated values for the *β* parameters which can provide a good fit with the available deaths data. To this aim, it is also important the role played by the coefficient of determination **r**^2^, which is a very informative parameter, needed to evaluate the goodness-of-fit of the simulated *Y* values (of the entire segment) versus the measured *Y* values (i.e., the available deaths data of Figure 1).

It is worth noticing also the motivation why we decided to compute the regression function in segments (i.e. pieces) which is based on the observation that COVID-19 deaths data follow different linear trends over the different periods (i.e., the colored sectors of Figure 1). Our segments will obviously result to be not connected, which is typical when the function to fit presents several alternating upward/downward oscillations.

Summarizing, our model will return the values of the *β*_*1*_ parameters for all segments, which, in turn, will be used to evaluate the variation of the steepness of the COVID-19 death trends for the seasons of interest. With the values of **r**^2^, finally, we will evaluate how well our segments fit with the available data.

The data presented in Figure 1, plus the code we developed to develop our segmented regression model, can be downloaded from: https://github.com/EugenioDeRosa/Covid-19_Linear_Regression.git. All the results of this study are fully reproducible by using the methods described in this Section, plus the data and the code available at the links above. Further reasonable requests relative to the data and the code can be also addressed to the corresponding author.

#### 2.3.1. Ethics approval of research

This study uses publicly available, aggregated data that contains no private information. Therefore, ethical approval is not required.

## 3. Results

Figures from 2 to 4 show the regression segments obtained with our piecewise regression model for all the winter, summer and intermediate periods of Figure 1.

Figure 2 is comprised of three different plots. From top to bottom the regression segments for: Winter 2021, Winter 2022, Winter 2023. Similarly for Figure 3. From top to bottom: Summer 2022, Summer 2023 and Summer 2024. Finally, Figure 4. From top to bottom: Intermediate 2022, Intermediate 2023 and Intermediate 2024.

**Figure 2.**
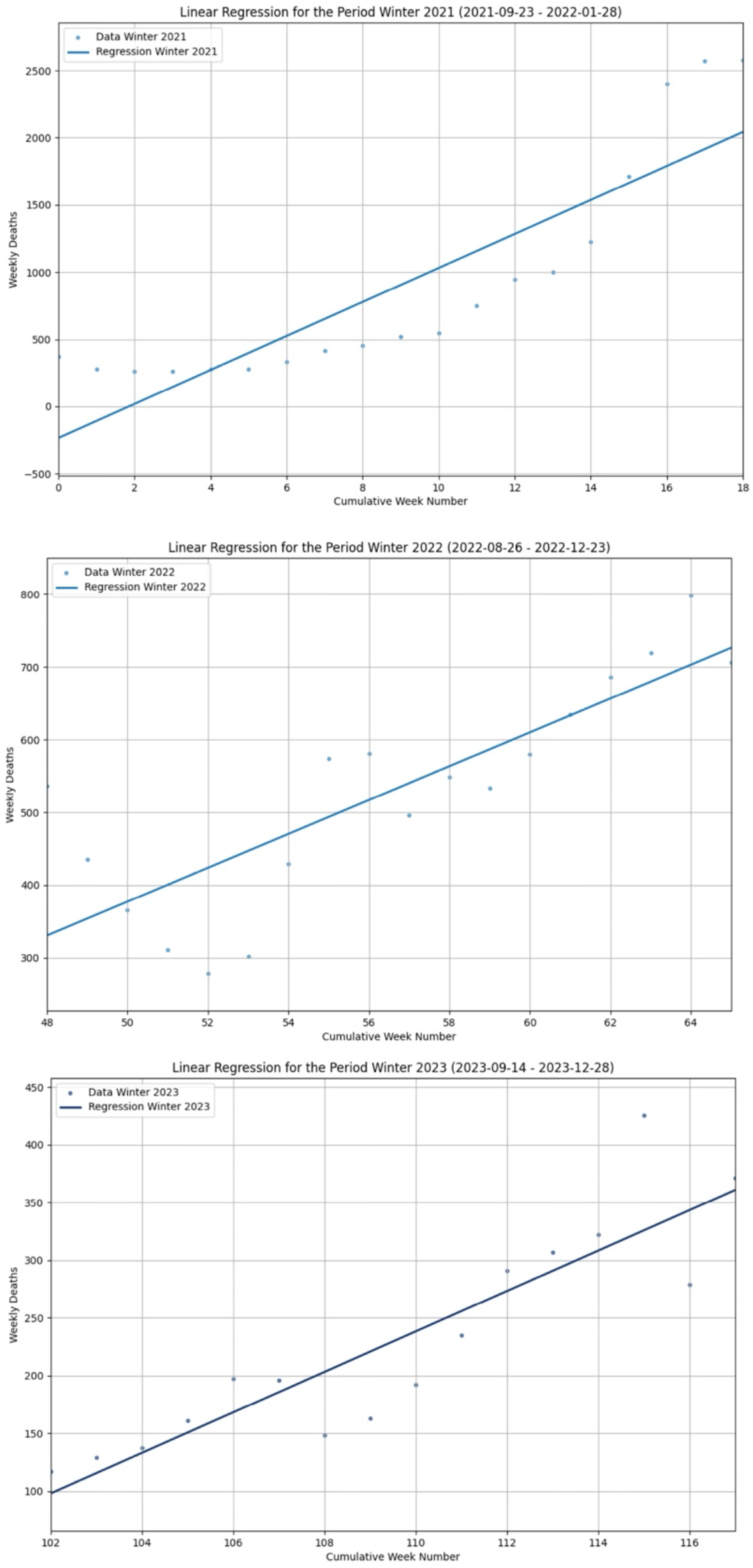
Regression segments for the Winter periods. Top: Winter 2021 (*β*_*1*_ =126.45, *β*_*0*_ = -233.70, **r**^2^ = 0.76). Middle: Winter 2022 (*β*_*1*_ = 23.28, *β*_*0*_ = 330.72, **r**^2^ = 0.67). Bottom: Winter 2023 (*β*_*1*_ = 17.52, *β*_*0*_ = 27.99, **r**^2^ = 0.80).

**Figure 3.**
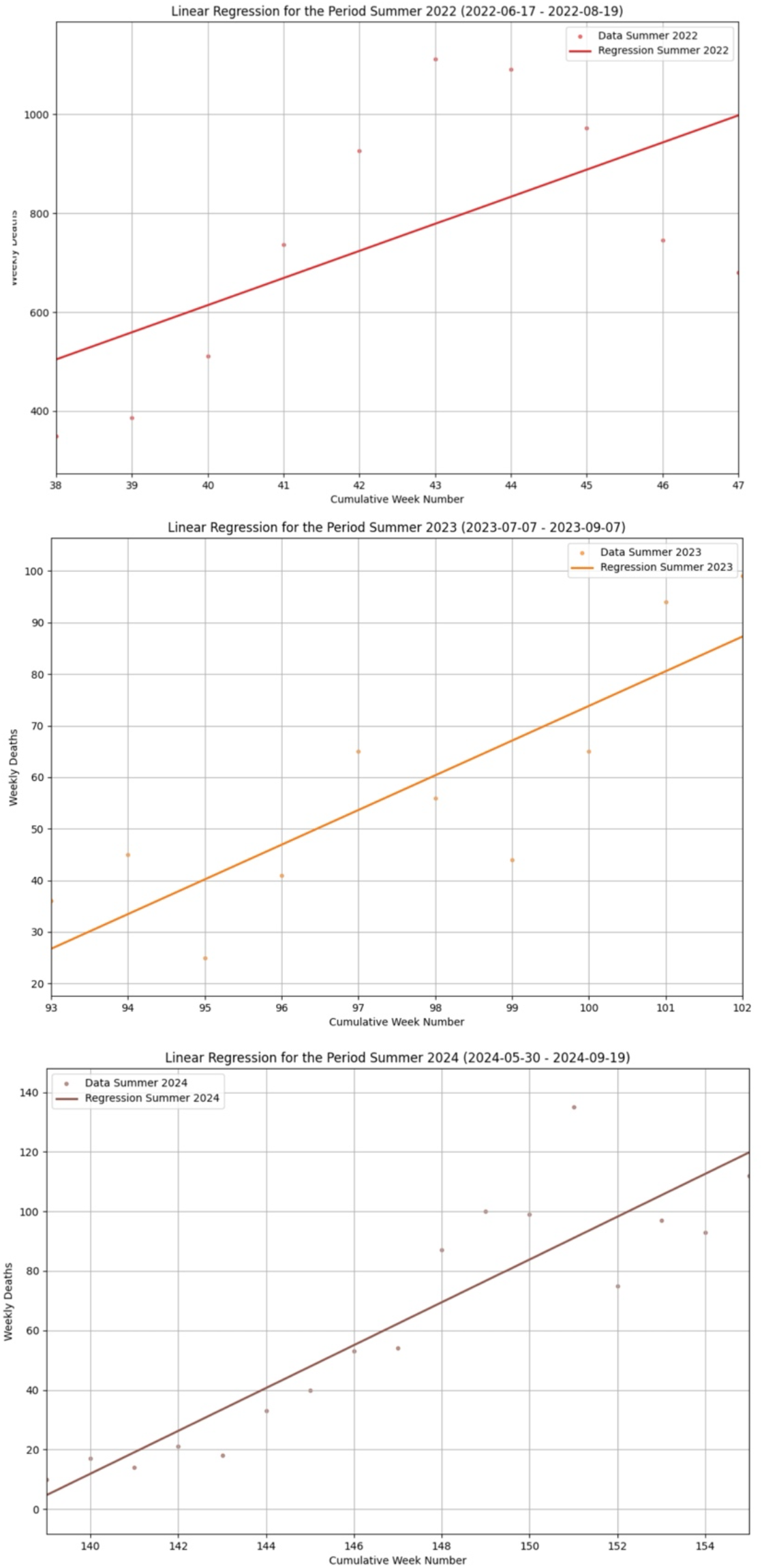
Regression segments for the Summer periods. Top: Summer 2022 (*β*_*1*_ = 54.80, *β*_*0*_ = 504.40, **r**^2^ = 0.36). Middle: Summer 2023 (*β*_*1*_ = 6.72, *β*_*0*_ = 26.72, **r**^2^ = 0.70). Bottom: Summer 2024 (*β*_*1*_ = 7.20, *β*_*0*_ = 4.70, **r**^2^ = 0.82).

**Figure 4.**
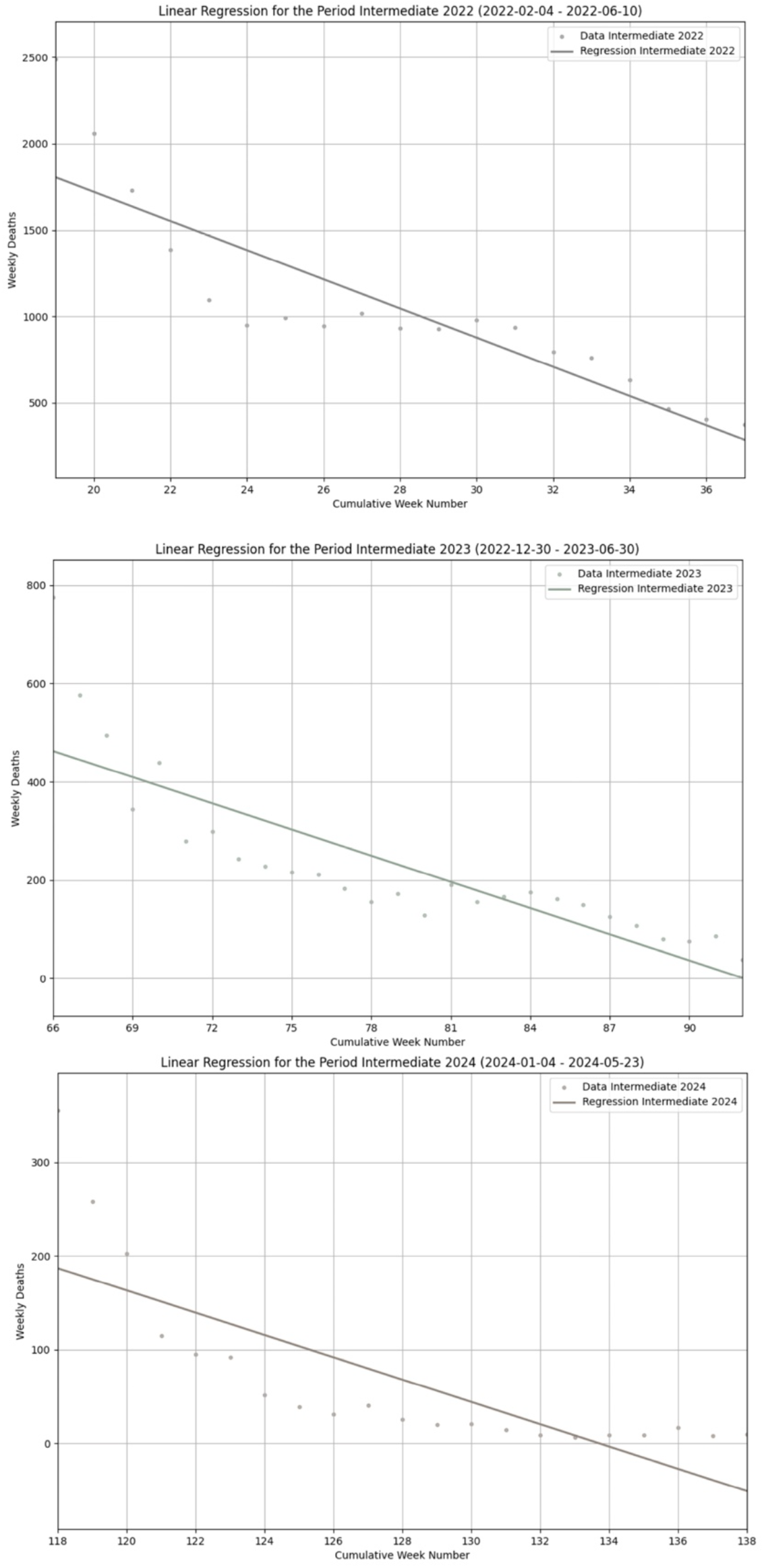
Regression segments for the Intermediate periods. Top: Intermediate 2022 (*β*_*1*_ = -84.38, *β*_*0*_ = 1805.09, **r**^2^ = 0.77). Middle: Intermediate 2023 (*β*_*1*_ = -17.76, *β*_*0*_ = 462.91, **r**^2^ = 0.71). Bottom: Intermediate 2024 (*β*_*1*_ = -11.89, *β*_*0*_ = 187.01, **r**^2^ = 0.62).

Each plot reports the dates of the beginning and the end of the considered periods. In all the plots, *Y* represents the number of weekly COVID-19 deaths for each given week registered along the *X* axis. Each Figure comes also with the values of the parameters *β*_*0*_, *β*_*1*_ and **r**^2^, computed for each single plot, specified in the corresponding captions. The measurements of the *β*_*1*_ and **r**^2^ parameters, per each different season, are also summarized in Table 2, along with the average and the standard deviation values, computed per each type of the different seasons under consideration.

**Table 2.** Characteristics of the regression segments for COVID-19 deaths of Figures 2-4: values of *β*_*1*_ and **r**^2^ per period, plus average and standard deviation per seasons.

| Period | $\beta_l$ | $r^2$ | $\beta_l$: Average,<br>and (standard<br>deviation) per<br>type of season | $r^2$: Average,<br>and (standard<br>deviation) per<br>type of season |
| --- | --- | --- | --- | --- |
| Winter 2021 | 126.45 | 0.76 | 55.75 (61.29) | 0.74 (0.05) |
| Winter. 2022 | 23.28 | 0.67 | 55.75 (61.29) | 0.74 (0.05) |
| Winter 2023 | 17.52 | 0.80 | 55.75 (61.29) | 0.74 (0.05) |
| Summer 2022 | 54.80 | 0.36 | 22.90 (22.55) | 0.63 (0.05) |
| Summer 2023 | 6.72 | 0.70 | 22.90 (22.55) | 0.63 (0.19) |
| Summer 2024 | 7.20 | 0.82 | 22.90 (22.55) | 0.63 (0.19) |
| Inter. 2022 | -84.38 | 0.77 | -38.01 (32.87) | 0.70 (0.19) |
| Inter. 2023 | -17.76 | 0.71 | -38.01 (32.87) | 0.70 (0.19) |
| Inter. 2024 | -11.89 | 0.62 | -38.01 (32.87) | 0.70 (0.06) |

As already anticipated, while the slope of a regression segment depicted in a given plot offers a visual impression of the velocity with which the corresponding COVID-19 mortality trend is increasing or decreasing, the numerical value of the associated *β*_*1*_ parameter provides the exact number by which COVID-19 deaths are progressively increasing with each new week of that period.

On the other end, **r**^2^ informs on how well a given linear regression segment has fitted the available observations (i.e., the initial COVID-19 deaths), in a scale from 0 to 1 (0-100%).

Several facts are noteworthy examining Figures 2-4 and Table 2. First, we can observe that: all the winter periods follow an increasing COVID-19 mortality trend, with positive slopes of the regression segments (Figure 2). Similarly, all the summer periods follow an increasing mortality trend (Figure 3). On the contrary, all the intermediate periods decline along a decreasing trend, with negative slopes of the corresponding regression segments (Figure 4).

Second, the increasing trends of winters and summers are different: the increasing mortality trend of winters is more pronounced, with an average *β*_*1*_ value of 55.75 versus an average *β*_*1*_ value for summers of 22.90 (fourth column of Table 2).

Third, there is an high variance of the slope coefficients within similar periods over different years, with a tendency towards less positive/negative slope coefficients with the passage of years.

Take Winters: we begin with a *β*_*1*_ value of 126.45 for Winter 2021, we proceed with a *β*_*1*_ value of 23.28 for Winter 2022, and we conclude with a *β*_*1*_ value of 17.52 for Winter 2023, yielding a SD value for *β*_*1*_ of 61.29 (second and fourth columns of Table 2).

This situation repeats similar during the summer periods, with the following values: Summer 2022 (*β*_*1*_ = 54.80), Summer 2023 (*β*_*1*_ = 6.72), Summer 2024 (*β*_*1*_ = 7.20). The SD value for *β*_*1*_ is equal to 22.55 (second and fourth columns of Table 2).

With intermediate periods, although negative, we observe the slopes becoming progressively less negative going from 2022 to 2024, with an average value of the *β*_*1*_ coefficient of -38.01 (SD 32.87), and a series of consecutive values (2022 - 2024) of *β*_*1*_ which are as follows: – 84.38, -17.76, -11.89 (second and fourth columns of Table 2).

Finally, if we consider the three Figure 2-4 as a whole, we can observe that the decreasing trends of the COVID-19 mortality, over all the three intermediate periods with their relatively long duration in time, have played the important role of compensating the upward drifts registered during winters and summers, thus contributing to the general declining trend of the COVID-19 mortality registered on the entire period of interest.

Coming to the **r**^2^ values, they confirm that our segmented linear model has a good fit with the initial COVID-19 deaths data (with just an exception). In fact, the winter periods (third and fifth columns of Table 2) show very good **r**^2^ values, precisely: Winter 2021 (**r**^2^ = 0.76), Winter 2022 (**r**^2^ = 0.67), Winter 2023 (**r**^2^ = 0.80), with an average value of **r**^2^ of 0.74 (SD 0.05). Similarly for the intermediate periods (third and fifth columns of Table 2): Intermediate 2022 (**r**^2^ = 0.77), Intermediate 2023 (**r**^2^ = 0.71), Intermediate 2024 (**r**^2^ = 0.62), with an average value of **r**^2^ of 0.70 (SD 0.06). Something slightly different happens for summers with the following values (third and fifth columns of Table 2): Summer 2022 (**r**^2^ = 0.36), Summer 2023 (**r**^2^ = 0.70), Summer 2024 (**r**^2^ = 0.82), yielding an average **r**^2^ value of 0.63 (SD 0.19).

Indeed, the low value of **r**^2^ for Summer 2022 is a consequence of what happened during that season when a rapid ascending trend of COVID-19 deaths was registered, beginning approximately at mid of June 2022, but peaking very soon on July 7, with 1,111 deaths, to return to its previous baseline values at mid of August [17]. Unfortunately, the rapid up and down of this summer pandemic wave has hardly a good fit with any linear model. Consequently, the slope of our regression segment, in this specific case, does not reflect well the speedy change from an upward to a downward drift of the COVID-19 mortality profile of those weeks, thus explaining the corresponding low value for **r**^2^ in this specific case. Regarding our segmented linear regression model, it is also worth noticing that all the *p-values* computed during the use of our model have been, to a large degree, below the significative value of *α* = 0.05, thus providing a further confirmation of the validity of this analysis.

To conclude this Section, we finally report in Figure 5 a comprehensive graphical summary of our results. They consist in the initial COVID-19 deaths data time series of Figure 1, divided over the nine different seasonal periods of interest, with superimposed the linear regression segments computed by our model and previously presented in Figures 2-4.

**Figure 5.**
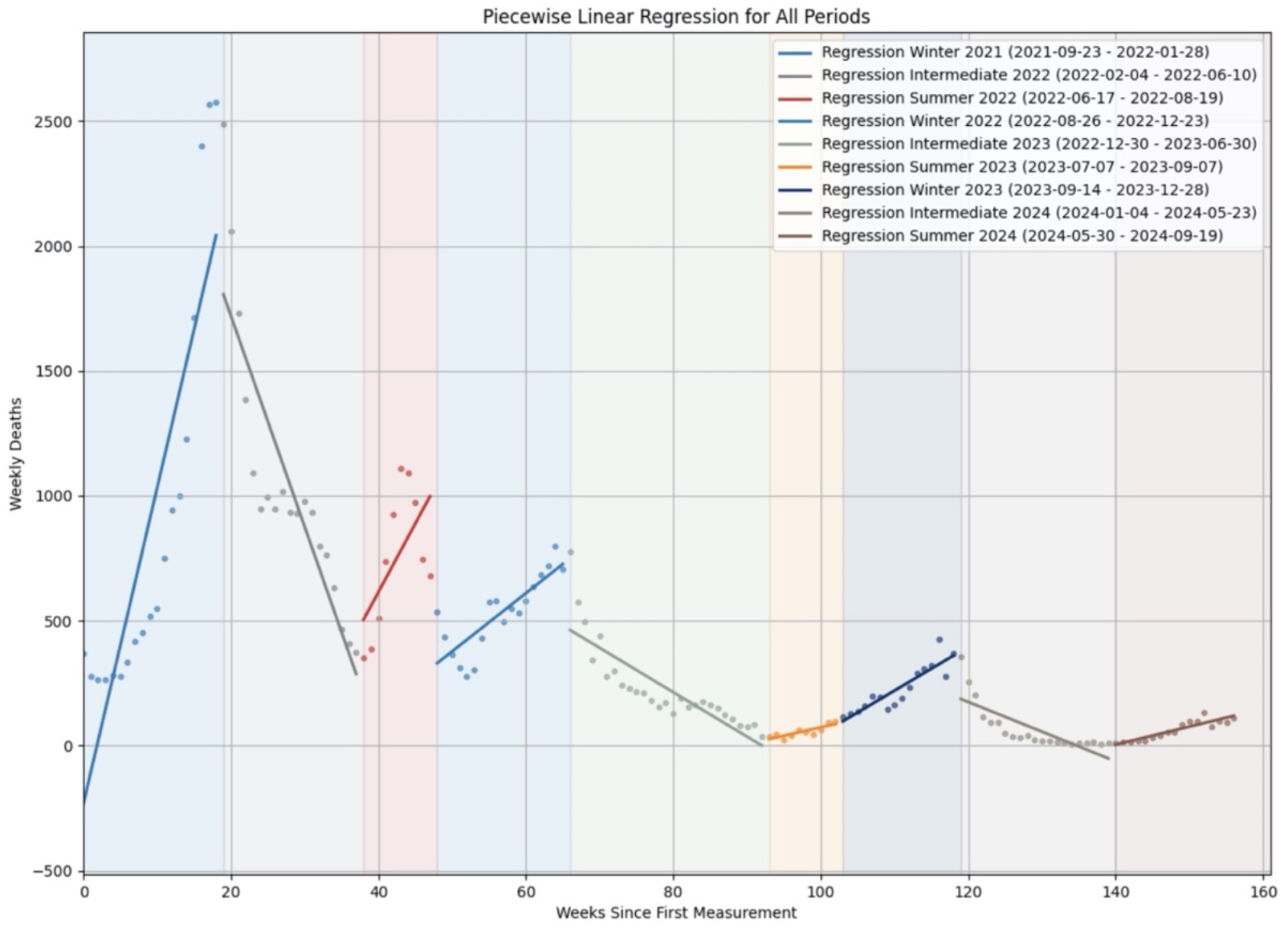
A comprehensive graphical summary of the results with the initial COVID-19 deaths data and superimposed the linear regression segments distributed over the nine different seasonal periods of interest.

## 4. Discussion

Previous studies have demonstrated that a strong evidence of a sinusoidal seasonal pattern that repeats over a one-year period cannot be found for the COVID-19 illness, at least in Western countries [10, 18, 19]. Nonetheless, with the present study, we have demonstrated that both ascending and descending seasonal COVID-19 mortality trends have been observed in Italy over all the period since September 2021 until September 2024.

In particular, the positive slopes of the segments of a piecewise linear regression model, fitted with the COVID-19 deaths data time series of that three-year long period, have revealed the recurrence of ascending COVID-19 mortality trends in all the winters and in the summers of that period, but more pronounced in winters.

Instead, the segments associated to all the three intermediate periods, extending from the end of winter to the beginning of summer, with their negative slopes, have revealed descending mortality trends that have played the role to compensate the upward drifts registered during winters and summers, thus contributing to the general decreasing rate of the COVID-19 mortality.

In the end, these seasonal upward/downward oscillations, repeated for three years, have contributed to go from an average number of weekly deaths from COVID-19 of almost 1000 in the period October 2021 - September 2022 to around 100 in the period October 2023 - September 2024 [3, 4].

All this said, the first limitation of our study is that it has scrutinized COVID-19 deaths data from a very specific time period (September 2021-September 2024). In that period, the epidemiological landscape in Italy was that of when the initial Omicron variant took over and then diversified into multiple post-Omicron sub-variants that gained an extremely increased survival fitness, leading to pandemic waves recurring in different seasons of the same year [20]. While the mathematical approach we have used to derive our findings remains valid, different results could be obtained examining different pandemic periods with the circulation of different SARS-COV-2 lineages.

We also recognize that this study has avoided identifying the motivations behind the upward/downward seasonal drifts we have identified. They can be attributable to several, different causes (or even to a combination of them), including: i) climatic and environmental factors, ii) social behaviors, like dense people gathering during holydays and vacations or common spreading events, iii) decreasing immunity from previous infections and vaccinations, and iv) various kinds of control and prevention measures. We are aware that further research is necessary to evaluate the role of those triggers and factors in the seasonal variations of mortality from COVID-19 [6].

Nonetheless, while this can be seen as a limitation of our research, we have decided to avoid taking part in the discussion about the causes of the seasonal COVID-19 deaths oscillations, with the precise idea to observe a natural phenomenon only to detect the presence of those seasonal increasing/decreasing mortality trends with neutrality, and regardless of the underlying factors.

Another characteristic of our study has been the decision to resort to a linear mathematical model (i.e., a piecewise linear regression model) resulting in a segment between two points. We are perfectly aware that this method is not the one with which COVID-19 deaths are usually counted [21, 22, 23]. Nonetheless, it should be clear that the target of our research was not trying to create a model for a standard count data analysis of COVID-19 deaths [24, 25, 26]. Rather, we were interested in looking at how quickly COVID-19 seasonal mortality trends grew or declined in a given season. In this sense, our linear regression model has been particularly useful in providing a clear identification of the COVID-19 mortality trend for each season of interest. It has also helped us to compare the slopes of different seasonal mortality profiles, over various years, showing the seasons to take under more control, at the level of health policy recommendations, for the benefit of that subset of at risk population who are seasonally vulnerable.

Similar arguments can be offered regarding the issue about the goodness-of-fit of our regression model with respect to the available COVID-19 deaths data. Again, it should be clear that we were not looking for the best-optimized model, but for a set of regression segments able to guarantee an acceptable approximation of the available deaths data, being acceptable any segment with a coefficient of determination above the threshold of 60/65%, as indicated in the specialized literature [27].

On the other end, using a segmented linear regression model has given the advantage of a temporal decomposition of the time series of the COVID-19 deaths data, allowing to differentiate between severe and moderate variations of the COVID-19 deaths trends, while distinguishing seasonal alterations from minimal upward/downward drifts of the time series. To conclude this discussion, we believe that, in our case, these technical limitations have touched more upon the specificity of the investigated topics rather than addressing a weakness of our analysis. Moreover, writing about them should help towards an in-depth understanding of these issues [28, 29, 30].

Limitations reside, finally, in the use of Italian data. In fact, on the one end, the extension to different geographies could result into different results. On the other end, we used data made available by the Italian Government under the form of aggregated measures from two different sources (Civil Protection Department and Ministry of Health). In several cases, those measures have changed value over time, subject to corrections and adjustments, reaching a relative stability only recently.

## 5. Conclusions

We have fitted a segmented linear regression model to investigate the occurrence of seasonal variations of COVID-19 mortality trends in the period September 2021 - September 2024, when the SARS-CoV-2 Omicron and post-Omicron variants were mostly predominant in Italy. The slopes of the segments of our regression model have suggested that, despite a general declining trend of the COVID-19 mortality over the entire period of observation, seasonal increasing variations of deaths from COVID-19 have occurred during all the winters and the summers of the period, but they were more pronounced in winters. Since these increasing seasonal alterations of the COVID-19 mortality have been always compensated by consistent downward drifts occurring during the intermediate periods between winters and summers, they have been considered cause for concern only occasionally. With our research, we have provided an important contribution for the benefit of that subset of at risk population who are seasonally vulnerable, having clearly identified the seasons to consider with more attention.

## Data Availability

All data produced in the present work are contained in the manuscript or downloadable from public data repositories

https://github.com/EugenioDeRosa/Covid-19_Linear_Regression.git

## Use of AI tools declaration

The authors declare that they have not used artificial intelligence (AI) tools in the creation of this article.

## Acknowledgments

This research received no external funding. The Authors are grateful to several colleagues from the University of Bologna who provided comments on a previous preprint version of this paper.

## Informed Consent Statement

Not applicable: Neither humans nor animals nor personal data are involved in this study.

## References

1. La Rosa G, Iaconelli M, Veneri C, et al, The rapid spread of SARS-COV-2 Omicron variant in Italy reflected early through wastewater surveillance. Science of The Total Environment, 2022, 837, doi: 10.1016/j.scitotenv.2022.155767

2. Bergami M, Bullini Orlandi F, Giuri P, et al, Embracing tensions throughout crises: The case of an Italian university hospital during the COVID-19 pandemic. Health Care Management Review, 2022, 49(3), doi: 10.1097/HMR.0000000000000404

3. Alicandro G, Gerli A, Remuzzi G, et al, Updated estimates of excess total mortality in Italy during the circulation of the BA.2 and BA.4-5 Omicron variants: April-July 2022. Work, Environmental and Health, 2022, 113(5), doi: 10.1097/HMR.0000000000000404

4. Mattiuzzi C, Lippi G, Nationwide analysis of COVID-19 death rate throughout the pandemic in Italy. Journal of Laboratory and Precision Medicine, 2023, 8(1), doi: 10.21037/jlpm-22-75

5. WHO – United Nations, WHO chief declares end to COVID-19 as a global health emergency. UN News, 2023, https://news.un.org/en/story/2023/05/1136367

6. Townsend JP, Hassler HB, Lamb AD, et al, Seasonality of endemic COVID-19. Journal of American Society for Microbiology, 2023, 14(6), doi: 10.1128/mbio.01426-23

7. Baum F, Freeman T, Musolino C, et al, Explaining COVID-19 performance: what factors might predict national responses? The BMJ, 2021, 372(91), doi: 10.1136/bmj.n91

8. Casini L, Roccetti M, Reopening Italy’s schools in September 2020: a Bayesian estimation of the change in the growth rate of new SARS-CoV-2 cases. BMJ Open, 2021, 11:e051458, doi: 10.1136/bmjopen-2021-051458

9. D’Amico M, Shaman J, Dubrow R, et al, COVID-19 seasonality in temperate countries, Environmental Research, 2022, 206(15), doi: 10.1016/j.envres.2021.112614

10. Cappi R, Casini L, Tosi D, et al, Questioning the seasonality of SARS-COV-2: a Fourier spectral analysis. BMJ Open, 2022, 12:e061602, doi: 10.1136/bmjopen-2022-061602

11. Chirumbolo S, Pandolfi S, Valdenassi L, Seasonality of COVID-19 deaths. Did social restrictions and vaccination actually impact the official reported dynamic of COVID-19 pandemic in Italy? Environmental Research, 2022, Volume 212, doi: 10.1016/j.envres.2022.11322

12. Taljaard M, McKenzie JE, Ramsay CR, et al, The use of segmented regression in analysing interrupted time series studies: an example in pre-hospital ambulance care. Implementation Science, 2014, 9(77), doi: 10.1186/1748-5908-9-77

13. Wiemken TL, Khan F, Puzniak L, et al, Seasonal trends in COVID-19 cases, hospitalizations, and mortality in the United States and Europe. Scientific Reports, 2023, Volume 13, doi: 10.1038/s41598-023-31057-1

14. Wong C, Why do covid cases rise in summer? New Scientist, 2024, 263(3506), doi: 10.1016/S0262-4079(24)01548-3

15. Zhang SX, Marioli A, Gao R, et al., A Second Wave? What Do People Mean by COVID Waves? – A Working Definition of Epidemic Waves. Risk Management and Healthcare Policy, 2021, Volume 14, doi: 10.2147/RMHP.S3260515

16. Altman N, Krzywinski M, Simple linear regression. Nature Methods, 2015, Volume 12, doi: 10.1038/nmeth.3627

17. Venturelli F, Mancuso P, Vicentini M, et al, High temperature, COVID-19, and mortality excess in the 2022 summer: a cohort study on data from Italian surveillances. Science of The Total Environment, 2023, Volume 887, doi: 10.1016/j.scitotenv.2023.164104

18. Fontal A, Bouma MJ, San-José A, et al, Climatic signatures in the different COVID-19 pandemic waves across both hemispheres, Nature Computational Science, 2021, Volume 1, doi: 10.1038/s43588-021-00136-6

19. Sera F, Armstrong B, Abbott S, et al, A cross-sectional analysis of meteorological factors and SARS-CoV-2 transmission in 409 cities across 26 countries. Nature Communications, 2021, 12(1), doi: 10.1038/s41467-021-25914-8

20. Stefanelli P, Trentini F, Petrone D, et al, Tracking the progressive spread of the SARS-CoV-2 Omicron variant in Italy, December 2021 to January 2022. Eurosurveillance, 2022, 27(45), doi: 10.2807/1560-7917.ES.2022.27.45.2200125

21. Khajanchi S, Sarkar K, Mondal J, et al., Mathematical modeling of the COVID-19 pandemic with intervention strategies. Results in Physics, 2021, Volume 25, doi: 10.1016/j.rinp.2021.104285

22. Hao B, Liu C, Wang Y, et al, A mathematical-adapted model to analyze the characteristics for the mortality of COVID-19. Scientific Reports, 2022, Volume 12, doi: 10.1038/s41598-022-09442-z

23. Roccetti M, Drawing a parallel between the trend of confirmed COVID-19 deaths in the winters of 2022/2023 and 2023/2024 in Italy, with a prediction. Mathematical Biosciences and Engineering, 2023, 21(3), doi: 10.3934/mbe.2024165

24. Tosi D, Campi A, How Data Analytics and Big Data Can Help Scientists in Managing COVID-19 Diffusion: Modeling Study to Predict the COVID-19 Diffusion in Italy and the Lombardy Region. Journal of Medical Internet Research, 2020, 22(10), doi: 10.2196/21081

25. Roccetti M, Excess mortality and COVID-19 deaths in Italy: A peak comparison study. Mathematical Biosciences and Engineering, 2023, 20(4), doi: 10.3934/mbe.2023304

26. Konstantinoudis G, Cameletti M, Gòmez-Rubio V, et al., Regional excess mortality during the 2020 COVID-19 pandemic in five European Countries. Nature Communications, 2022, 13(1), doi: 10.1038/s.41467-022-28157-3

27. Ozili PK, The acceptable R-square in empirical modelling for social science research. 2022, Social Research Methodology and Publishing Results, doi: 10.2139/ssrn.4128165

28. Corradini F, Gorrieri R, Roccetti M, Performance preorder and competitive equivalence. Acta Informatica, 1997, 34(11), doi: 10.1007/s002360050107

29. Palazzi CE, Ferretti S, Cacciaguerra S, et al, Interactivity-loss avoidance in event delivery synchronization for mirrored game architectures. IEEE Transactions on Multimedia, 2006, 8(4), doi: 10.1109/TMM.2006.876229

30. Agosto A, Campmas A, Giudici P, et al, Monitoring COVID-19 contagion growth. Statistics in Medicine, 2021, 40(18), doi: 10.1002/sim.9020

